# Predicting Rates of Cognitive Decline in Older Adults with Brain- and Cardiac-Imputed Transcripts

**DOI:** 10.64898/2026.09.29.26364189

**Authors:** Ali Razavi, Nathan R. Tucker, Auyon J. Ghosh, Stephen V. Faraone, Stephen J. Glatt, Jonathan L. Hess

## Abstract

Severe cognitive decline due to dementia is an important clinical indicator signaling neurodegeneration. Predicting cognitive decline rates can be useful in establishing prognosis and modifying treatment regimens. The rate of decline is highly variable across patients with dementia and may be determined by an interplay of many factors. This variability poses challenges to existing tools for predicting disease progression. Changes in heart and brain tissue have been associated with cognitive decline, yet tissue-specific molecular data are difficult to obtain. In this study, we sought to estimate cognitive decline-related transcripts in the brain and heart of people actively living with and without dementia. We reconstructed transcriptomic profiles in brain and heart with the transcriptome-imputation algorithms *BrainGENIE* and *HeartGENIE,* and identified several hundreds of transcripts associated with cognitive decline over a two-year assessment window, with the cerebellar hemisphere, left ventricle of the heart, and atrial appendage showing the largest number of cognitive-related transcripts. Pathway analysis revealed an enrichment among cognition-related transcripts of gene sets involved in synaptic, proteostasis/autophagy, immunologic, and mitochondrial respiratory chain functioning. Hundreds of perturbagens show significant concordance with cognitive decline-related transcriptomic signatures in brain and heart, pointing to potentially druggable targets. Top brain- and heart-specific cognition-associated transcripts were incorporated into LASSO regression models predicting future cognitive decline and were compared to two models, one utilizing age, sex and educational attainment, and a hybrid model incorporating both data types. Our models were able to achieve statistically significant correlations in the test set, yet had weak overall. Together, these findings demonstrate that brain and heart transcriptomes capture distinct aspects of biology related to cognitive decline, providing more nuanced insights into normal and aberrant brain aging that may inform drug-repurposing and risk-prediction efforts.

## INTRODUCTION

Dementia is one of the leading causes of disability in the elderly and Alzheimer’s Disease (AD) is the leading cause of dementia with over 6 million affected individuals in the US (Alzheimer’s Association, 2025). It is a progressive and fatal disorder with limited treatment options. The hallmark of AD is progressive cognitive decline, a symptom shared by other types of dementia.

Cognitive decline rates vary across individuals and have consequences in disease profiling, prognosis and treatment regimens. Cognitive decline prediction can be useful in establishing prognosis and modifying treatment regimens as: (1) it enables the detection of individuals at risk for rapid deterioration, which aids families and healthcare providers in better preparing for care; (2) available and future AD treatments have been predicted to have better efficacy when started early in the disease process (Bregman et al., 2025) (Pais et al., 2020 and (3) variables that can predict cognitive decline rates could elucidate the underlying pathophysiology of dementia.

The molecular basis of cognitive decline has been primarily studied through two complementary approaches. Postmortem brain studies have identified molecular abnormalities in those who declined cognitively, but these changes are observed after substantial neurodegeneration had taken place (Johnson et al., 2022; Wingo et al., 2022; Mostafavi et al., 2018). Select markers nominated through postmortem brain studies can be subsequently tracked in plasma as dementia sets in and corroborated by neuroimaging (Cicognola et al., 2021; Nyberg et al., 2020; Chatterjee et al., 2022; Stevenson-Hoare et al., 2023). Plasma profiling has also nominated markers originating outside of the brain linked to cognitive decline (Lindbohm et al., 2022; Klinger et al., 2026; Adegboye et al., 2025; Bezgin et al., 2025; Madsen et al., 2015). However, the molecular states in the living human brain that precede and predict future cognitive decline remain largely unknown. Importantly, extensive epidemiological and genetic evidence points to a biological connection between the heart and brain, suggesting their interaction supports cognitive functioning. A myriad of cardiovascular diseases including hypertension, atherosclerosis, coronary heart disease, atrial fibrillation, obesity, diabetes, and vascular dysregulation have been associated with Alzheimer’s disease and cognitive decline (Livingston et al., 2020; Gorelick et al., 2011; Iturria-Medina et al., 2016; Debette & Seshadri, 2011). This raises the possibility that cardiac-specific molecular states could provide a complementary window into biological processes relevant to cognitive decline, potentially capturing systemic or heart-brain dependencies that are not readily accessible through brain-derived measures alone. Thus, tracking the molecular origins of cognitive decline in brain and heart can be important for a better understanding of dementia pathophysiology.

Numerous pathological processes have been associated with AD with varying degrees of empirical evidence, with directly implicated changes in the brain as well as peripheral tissues associated with decline. A large-scale meta-analysis of postmortem brain data in AD cases established dysregulation across many genes and pathways across cortical and subcortical regions (Hou et al., 2024). Nevertheless, the molecular states in the living human brain that precede cognitive decline remain difficult to characterize as living brain tissue is largely inaccessible, while the molecular changes linking the heart to cognition remain poorly understood. To close this knowledge gap, we utilized our transcriptome-imputation algorithms *BrainGENIE* and *HeartGENIE* to predict multi-organ gene-expression profiles within brain and heart *via* whole blood transcriptomic data (Hess et al.,2023) derived from older adults with and without dementia. Our goal was to identify molecular signatures in the brain and heart linked to future cognitive decline.

Our transcriptome-imputation models offer the advantage of being a proxy for tissue data without the need for direct biopsies and/or imaging data, as well as uncovering molecular-level associations with cognitive decline. Moreover, this approach offers the possibility of discovering antemortem regional molecular associations with cognitive decline. The increased molecular resolution offered via our model helps with accessible cognitive prediction and the generation of novel pathophysiological hypotheses, which can increase access to endpoints for clinical trials and opportunities for treatment and care optimization for providers and patients.

## METHODS

### Imputation of Transcripts in Brain and Heart

We accessed blood-based transcriptome-wide gene-expression profiles from study participants collected during baseline study visits. These data were generated in Religious Orders Study and Rush Memory and Aging Project (**ROSMAP**), AddNeuroMed, and Alzheimer’s Disease Neuroimaging Initiative (**ADNI**) cohorts, and were accessed with appropriate approvals. For ROSMAP and AddNeuroMed, non-normalized gene-expression data were available, to which we applied standard pre-processing methods, as previously described.^1^ Data-processing procedures are highlighted in the reference manuscript (Hou et al., 2025). Genes including protein-coding genes, miRNAs, and lncRNAs were kept with CPM ≥ 1 in at least 50% samples. The data were quantile-normalized with the *limma* package. Batch correction of normalized log_2_CPM values was performed using the *ComBat* algorithm in the *R* package *sva*.

AddNeuroMed transcriptome data were generated in two batches using Illumina Human HT-12 V3.0 Beadchip arrays. We applied log_2_ transformation and quantile-normalization to each batch, followed by median summarization of normalized probe intensities to obtain a single measure of expression per gene. Then we applied batch-correction using the *ComBat* algorithm in the *sva* library in *R*.

For study participants in ADNI, only normalized gene-expression data were available, which we accessed from the ADNI database (adni.loni.usc.edu). ADNI generated transcriptome-wide gene expression readouts using the Affymetrix U219 array, and applied the robust multi-array average (RMA) method and quantile normalization. Additional quality control steps are available on the ADNI website.

The *BrainGENIE^2^* and *HeartGENIE* transcriptome-imputation models were trained using pre-processed, normalized RNA-sequencing data from donors collected in the Genotype Tissue Expression (GTEx) project, version 10. These models are trained on GTEx v10 and predict gene-expression levels in 10 brain regions, along with the left ventricle and right atrial appendage of the heart. Documentation for the model architecture can be found in the reference paper (Hess et al., 2023), but in brief, the *GENIE* models are predicated on principal components of gene expression measured in peripheral blood, which are then used to impute the levels of expression of individuals transcript in the target tissue.

Models for significantly imputable transcripts were deployed on donor data from ROSMAP, AddNeuroMed, and ADNI to impute brain-regional and cardiac-specific transcript-expression levels in the following regions: amygdala, anterior cingulate cortex (BA24), caudate, cerebellar hemisphere, frontal cortex (BA9), hippocampus, hypothalamus, nucleus accumbens, putamen, substantia nigra, heart atrial appendage, and heart left ventricle.

### Measuring Cognition and Rates of Cognitive Decline Longitudinally

In each cohort, we derived person-specific slopes capturing the rate of decline in cognition based on repeat assessments of the Mini-Mental Status Exam (**MMSE**) over a two-year period. MMSE is an 11-question test of global cognitive performance reflected in a 30-point scale, with scores lower than 24 suggesting cognitive impairment. Rate of cognitive decline over multiple assessments is a frequently used endpoint for clinical trials.

Participants completed annual MMSE assessments in the ROSMAP and AddNeuroMed cohorts, whereas the ADNI cohort reevaluated their participants at months 3, 6, 12, 18, and 24 post-baseline.

We quantified person-specific slopes denoting the rate of cognitive decline of a two-year period *via* random-effect regression modeling with the *lmerTest* library in *R*. The regression formula was expressed as: MMSE_ij_=(β_0_+b_0i_)+(β_1_+b_1i_)⋅time_ij_+ε_ij_, which allowed both slope *<u>and</u>* intercept of the underlying effect of time on MMSE to vary by person.

### Transcripts Associated with Cognitive Decline

We sought to identify brain-regional and heart transcriptional states at baseline related to person-specific slopes reflecting rate of cognitive decline. AddNeuroMed cohort data were chosen for this analysis due to a larger sample size. Mixed-effect regression models were used with brain- and cardiac-imputed transcripts as the predictor variables and individual-level rates of cognitive decline in MMSE based on a two-year window of repeat cognitive assessments as the predicted variable. We used the *limma* package in *R* to fit the regression models which included participant age at the baseline study visit, biological sex, and years of education attained as covariates. Moderated *t*-statistics of the coefficient of MMSE decline slope in two years for each gene were calculated using the empirical Bayes method from the *eBayes* function. *P*-values were adjusted using the Bonferroni procedure to account for multiple comparisons for all statistical tests reported in the manuscript.

### Pathway Analysis

Pathway enrichment analyses were conducted on the *t*-statistics of the genes derived from the fitted MMSE model. We utilized the *fGSEA* package with the following settings: minSize=10, maxSize=2000, eps=0. Gene-set annotations were obtained from the *Reactome* knowledgebase (Jessal et al., 2020). For reporting, pathway-level FDR thresholds were set at ≤0.05.

To enhance the interpretability of the enrichment patterns, we summarized the pathway analysis results into 29 distinct *Reactome* categories (*Reactome* release 88, accessed 13 June 2025). These categories were defined according to the established hierarchical relationships within the *Reactome* database, represented by child–parent relationships.

### Connectivity Map (CMap) Drug Signatures

We performed a computational analysis to assess the druggability of cognition-related transcriptional states. *In vitro* perturbation profiles were obtained *via* Connectivity Map (CMap) for FDA-approved compounds; perturbation profiles represent transcriptional shifts in expression levels induced by each compound. Using Pearson’s correlation tests, we estimated the concordance between CMap’s perturbagen profiles and the cognition-transcript associations identified in our study. A significance threshold of *p*<0.05 after Bonferroni correction and a correlation value (*r*) above 0.1 or below −0.1 was chosen to identify compounds significantly correlated with cognitive decline signature in each imputed cardiac and brain region.

### Training, Cross-Validation, and External Testing of Cognitive Decline Prediction Models

We used the *R* library *cv.glmnet* to train Least Absolute Shrinkage and Selection Operator(LASSO) regression models with the aim of predicting the rate of MMSE change over a two-year assessment window based on the brain-regional- and cardiac-imputed transcripts predicted by *BrainGENIE* and *HeartGENIE* at the initial testing visit. These features were used alongside participants’ age at the initial testing visit and their sex assigned at birth. The purpose of this simplified model was to determine the predictive validity of imputed transcripts; not necessarily to maximize the amount of explained-varianced in or derive a best predictor of cognitive change over time.

The AddNeuroMed cohort was used to estimate prediction performance of the LASSO regression models through a single 5-fold cross-validation. The cross-validation framework was set up as follows: participants were randomly assigned into one of five equally sized validation folds. In each iteration of cross-validation, participants in one of the validation folds were set aside and we used the remaining participants for model training and optimization. In the training subset, we performed a transcriptome-wide differential expression analysis using *limma* to rank brain-regional- and cardiac-imputed transcripts according to their association with cognitive decline. These differential expression models included participants’ age at the testing visit and sex. The top 100 transcripts, ranked by significance of association with MMSE change over time, were entered into a LASSO regression model, along with age and sex, which further selected the most informative transcripts and demographic features to predict the individualized slope of MMSE decline. The LASSO regression was fit on our training subset with the *cv.glmnet* function, where the lambda parameter was tuned *via* nested 10-fold cross-validation to identify the minimum penalization value that minimizes mean-squared error of predicted *vs*. actual values for slope of MMSE decline. The model with the best L1 lambda value was applied to the participants in the withheld validation fold. A Pearson’s *r* correlation test was used to calculate the relationship between predicted *vs*. actual slope of MMSE decline, which was then averaged over the five validation folds to obtain a robust estimate of prediction accuracy. The Pearson’s *r* values from all validation folds were converted to *z*-scores *via* Fisher’s transformations, and resulting *z*-scores were used to calculate two-tailed *p*-values representing the significance of cross-validation prediction accuracy.

After establishing an estimate of prediction accuracy for this simplified “omics” model (*i.e.,* imputed transcripts + age + sex), we then sought to determine whether imputed transcriptional states at baseline enhance the predictability of MMSE change over time above that of models that rely upon variables related to basic demographics and years of education (as a proxy of cognitive reserve, a well-reported predictor of cognitive decline and dementia risk^5–7^). Hence, we performed a single 5-fold cross-validation analysis, as above, but restricted the LASSO regression models to using baseline age, sex, and years of education (up to baseline visit) as variables to predict slope of MMSE decline. We repeated the same process of tuning L1 penalization *via* nested 10-fold cross-validation, taking the mean Pearson’s *r* of predicted *vs*. actual slope of MMSE change over the 5 validation folds, and calculating *p*-values of prediction accuracy after converting fold-level Pearson’s *r* values to Fisher’s *z*-scores. This served as our simplified “clinical” model. Next, we integrated the best-performing features from our omics and clinical models into a “unified” model (*i.e.,* age + sex + years of education + brain-regional & cardiac-imputed transcripts), which was trained and optimized *via* a single 5-fold cross-validation. The same procedures were followed to calculate prediction accuracy.

We optimized each of the three models (“omics”, “clinical”, and “unified”) by tuning their parameters on the entire AddNeuroMed cohort. Finally, we applied each of the models to the ROSMAP and ADNI cohorts to evaluate their external generalizability. Prediction accuracy was evaluated using Pearson’s *r* tests comparing predicted *vs.* actual slope of MMSE decline.

## RESULTS

We compiled clinical as well whole blood gene expression data from 3 studies focused on cognitive decline. The total number of cases and controls in the threedatasets are as follows s: ROSMAP comprises 221 controls and 52 cases; AddNeuroMed is comprised of 191 controls and 377 cases; ADNI comprises 137 controls and 331 cases. A summary of demographic and clinical variables, along with participants’ cognitive decline status from each cohort is provided in **Table 1**.

**Table 1A.** A summary of the demographic characteristics of ROSMAP participants.

| Characteristic | CU<br>N = 221 | MCI<br>N = 45 | MCI+ other<br>impairment*<br>N = 1 | AD<br>N = 3 | AD+ other impairment<br>N = 3 |
| --- | --- | --- | --- | --- | --- |
| Sex (Males, %) | 47 (21%) | 8 (18%) | 0 (0%) | 3 (100%) | 2 (67%) |
| Age at blood sampling | 83.7 ± 0.4 | 84.9 ± 0.8 | 89.4 ± 0 | 82.6 ± 2.4 | 81.8 ± 6.2 |
| Years of education | 15.2 ± 0.2 | 15.4 ± 0.4 | 22.0 ± 0 | 17.0 ± 1.0 | 16.0 ± 2.0 |
| Baseline MMSE | 28.7 ± 0.1 | 27.6 ± 0.2 | 28.0 ± 0 | 10.0 ± 1.0 | 13.2 ± 5.0 |
| MMSE slope | 0.0 ± 0.0 | -0.1 ± 0.0 | -0.1 ± 0 | -0.2 ± 0.0 | -0.2 ± 0.0 |

\*Other impairment = other condition(s) related to cognitive impairment recorded in the ROSMAP dataset.

**Table 1B.** A summary of the demographic characteristics of AddNeuroMed participants.

| Characteristic | CU<br>N = 191 | MCI<br>N = 184 | AD<br>N = 193 |
| --- | --- | --- | --- |
| Sex (Males, %) | 76 (40%) | 80 (43%) | 60 (31%) |
| Age at blood sampling | 73.7 ± 0.5 | 74.9 ± 0.4 | 76.5 ± 0.5 |
| Years of education | 10.5 ± 0.3 | 9.4 ± 0.3 | 8.2 ± 0.3 |
| Baseline MMSE | 28.7 ± 0.2 | 27.1 ± 0.1 | 20.5 ± 0.3 |
| MMSE slope | 0.0 ± 0.0 | -0.1 ± 0.0 | -0.2 ± 0.0 |

**Table 1C.** A summary of the demographic characteristics of ADNI participants.

| Characteristic | CU<br>N = 137 | EMCI*<br>N = 208 | LMCI*<br>N = 83 | AD<br>N = 40 |
| --- | --- | --- | --- | --- |
| Sex (Males, %) | 64 (47%) | 112 (54%) | 43 (52%) | 24 (60%) |
| Age at blood sampling | 73.6 ± 0.5 | 71.0 ± 0.5 | 72.2 ± 0.8 | 74.8 ± 1.5 |
| Years of education | 16.6 ± 0.2 | 15.9 ± 0.2 | 16.8 ± 0.3 | 15.9 ± 0.4 |
| Baseline MMSE | 29.0 ± 0.1 | 28.3 ± 0.1 | 27.6 ± 0.2 | 22.9 ± 0.3 |
| MMSE slope | -0.01 ± 0.00 | -0.03 ± 0.00 | -0.1 ± 0.0 | -0.2 ± 0.0 |

\* EMCI = early mild cognitive impairment, LMCI= late mild cognitive impairment

### Regional Brain and Heart Tissue Imputation

To measure gene expression profiles among inaccessible brain and heart, we applied *HeartGENIE* and *BrainGENIE* to blood gene expression data from the ADNI, ROSMAP and AddNeuroMed cohorts which longitudinally tracked cognition in cognitively unimpaired and participants with dementia. imputed gene expression data in 10 brain regions and structures from two chambers of the heart. The number of significantly imputed transcripts at a *p*-value below 0.05 after FDR correction was as follows: right atrial appendage (*n*_genes_ = 18,083), anterior left ventricle of the heart (*n*_genes_ = 15,299), frontal cortex (*n*_genes_ = 11,278), cerebellar hemisphere (*n*_genes_ = 9,736), hippocampus (*n*_genes_ = 8,736), anterior cingulate cortex (*n*_genes_ = 8,020), amygdala (*n*_genes_ = 7,209), putamen (*n*_genes_ = 7,001), nucleus accumbens (*n*_genes_ = 8,845), caudate (*n*_genes_ = 10,477), substantia nigra (*n*_genes_ = 11,473), and hypothalamus (*n*_genes_ = 5,871).

### Brain-Regional and Cardiac Transcripts Associated with Cognitive Decline

Differential expression analysis revealed several hundreds of transcripts in heart and barin whose expression levels at baseline were significantly associated (Bonferroni *p*s < 0.05) with the pace of cognitive decline over the next two years (**Figure 1**). Cognition-related transcripts were identified in all 12 tissues. The cerebellar hemisphere exhibited the largest number of cognition-associated transcripts (225), closely followed by the left ventricle of the heart (149), and right atrial appendage (113). Additionally, cognition-related transcripts were found in the substantia nigra (61), caudate (38), nucleus accumbens (27), amygdala (26), and anterior cingulate cortex (25), hypothalamus (16), hippocampus (15), putamen (15), and frontal cortex (12).

**Figure 1.**
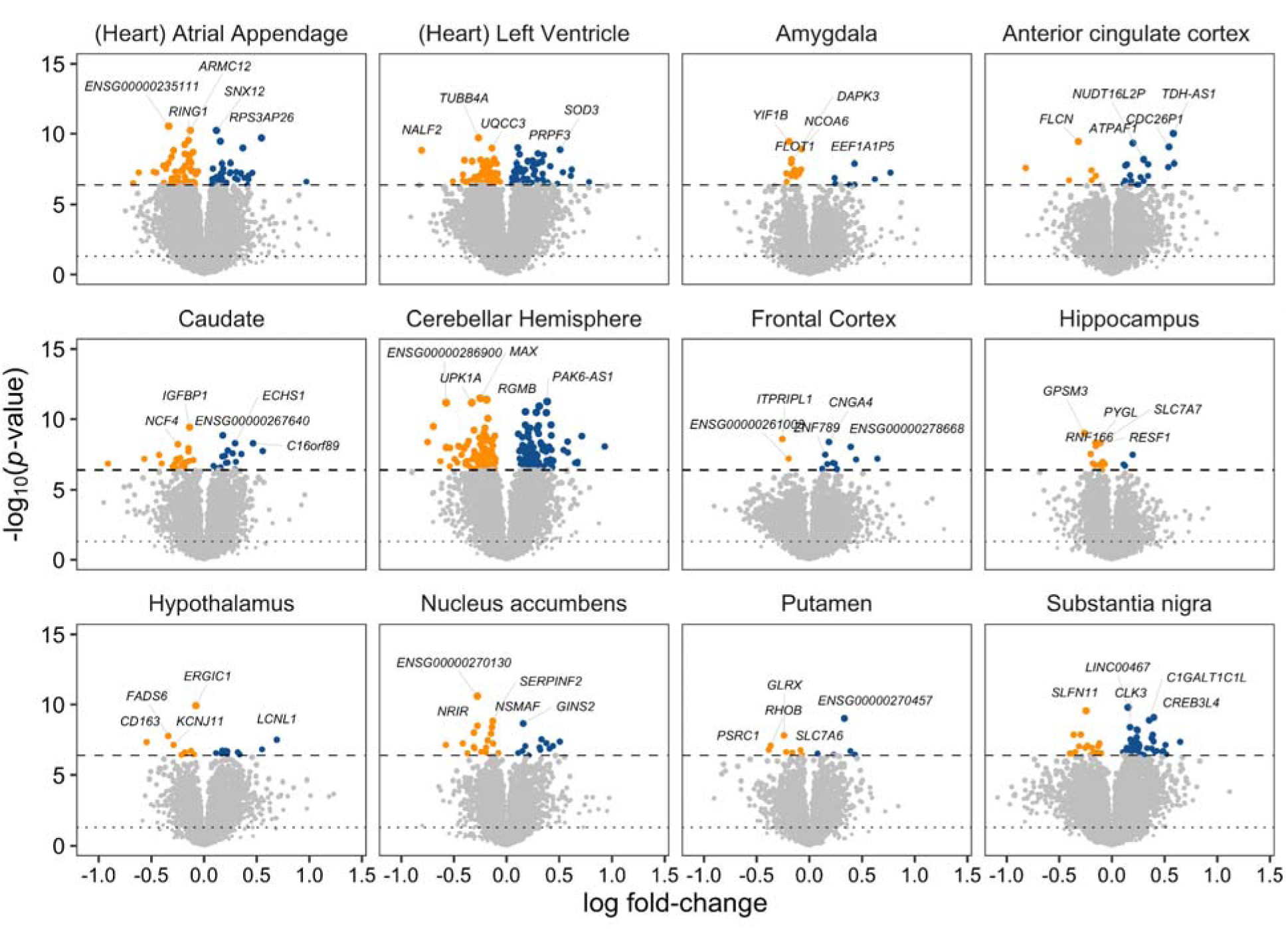
Volcano plots showing the association between MMSE change over time and baseline expression levels of brain-regional- and cardiac-imputed transcripts. The top ranking differentially expressed genes associated with cognitive decline are labeled in each panel. A dashed line denotes the Bonferroni-adjusted significance threshold, and a dotted line denotes an uncorrected *p* = 0.05. Transcripts corresponding to either pseudogenes or genes that have yet to be characterized appear with an Ensembl (“ENSG”) gene identifier.

Transcripts in the cerebellar hemisphere that were most prominently associated with cognition included *RGMB*, *UPK1A*, *MAX*, *PAK6-AS1*, and *MED29* (|t| > 7, FDR-adjusted *p*-value = 1.1×10). The top emergent hit from the heart was the transcript *RP1-29C18.8* from the right atrial appendage (*t* = −7.04, FDR-adjusted *p*-value = 1.1×10). The top cognition-related transcripts identified in the 12 tissues are provided in **Table 2** with a full set of results from our differential expression analyses available in **Supplementary Table 1**.

### Pathway Enrichment Analyses and Root Pathway Analysis

Gene set enrichment analyses identified dysregulated pathways in different tissues: 1,410 in the right atrial appendage of the heart, 1,140 in the left ventricle of the heart, 933 in the frontal cortex, 868 in the substantia nigra, 569 in the anterior cingulate cortex, 337 in the hippocampus, 238 in the cerebellar hemisphere, 107 in the amygdala, 79 in the hypothalamus, 41 in the nucleus accumbens, 36 in the caudate, and 30 in the putamen. Pathways consistently downregulated across tissues included metabolism, protein metabolism, cellular response to stimuli and disease. Pathways consistently upregulated across tissues included signal transduction, immune, chromatin organization and disease. The complete tissue-level pathway enrichment results are available in **Supplementary Table 2**. A total of 27 root pathways across the 12 imputed regions were dysregulated. Major patterns in the results included a consistent downregulation in the root pathways in the Heart Atrial Appendage and Heart Left Ventricular imputed tissue as well as major upregulation in the frontal cortex. The complete root pathway analysis results are reported in **Figure 2**.

**Figure 2.**
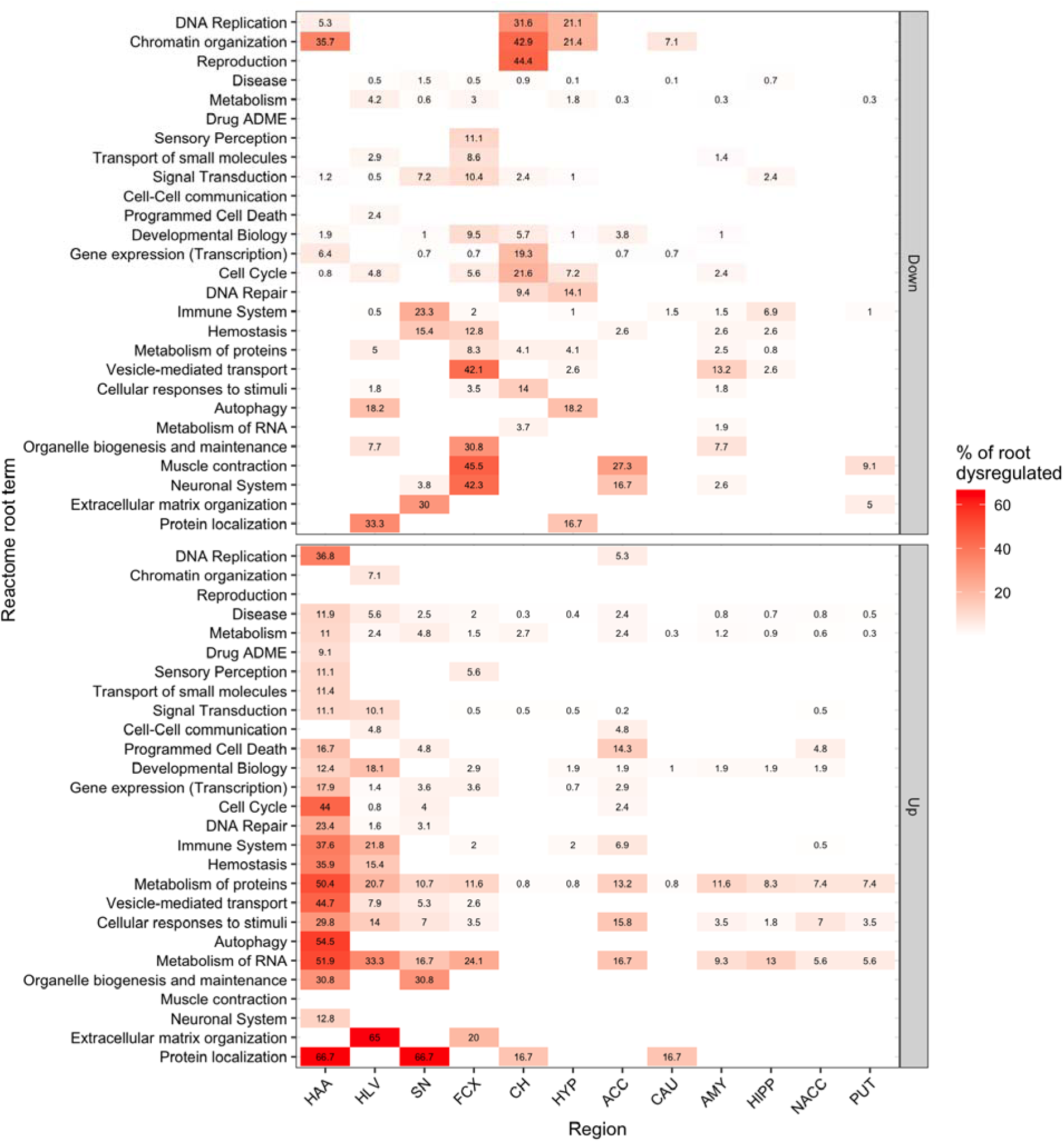
Hierarchically clustered heatmap of root pathway analysis of upregulated and downregulated pathways associated with cognitive decline. Each cell indicates the proportion of pathways within a given Reactome root category (nominal *p* < 0.05). Rows correspond to root Reactome categories. Columns correspond to the brain regions collected from GTEx donors and profiled by RNA-sequencing. Axes were hierarchically clustered (Euclidean distance, average linkage) on the underlying percent-coverage matrix so that adjacent rows and columns share similar enrichment profiles, facilitating visual comparison. The following are the abbreviations for the 2 heart regions and 10 brain regions: HAA: Heart Atrial Appendage, HLV: Heart Left Ventricle, AMY = Amygdala; ACC = Anterior cingulate cortex (BA24); CAU = Caudate nucleus (basal ganglia); CH = Cerebellar Hemisphere; FCX = Frontal Cortex (BA9); HIPP = Hippocampus; HYP = Hypothalamus; NACC = Nucleus accumbens (basal ganglia); PUT = Putamen (basal ganglia); SN = Substantia nigra

### Connectivity Map (CMap) Drug Signatures

We identified hundreds of known perturbagens that induce a transcriptomic shift that is significantly correlated with the expression-related signatures in brain and heart that we linked to cognition (Bonferroni-significant *ps* < 0.05). The correlations between the perturbagens and cognitive signatures exceeded an absolute value of 0.1. The total number of significant drug associations was as follows: right atrial appendage (*n*_perturbagens_ = 115), anterior left ventricle (*n*_perturbagens_ = 20), frontal cortex (*n*_perturbagens_ = 10), substantia nigra (*n*_perturbagens_ = 15), cerebellar hemisphere (N = 10), hippocampus (*n*_perturbagens_ = 10), anterior cingulate cortex (*n*_perturbagens_ = 10), caudate (*n*_perturbagens_ = 10), nucleus accumbens (*n*_perturbagens_ = 10), putamen (N = 5), amygdala (*n*_perturbagens_ = 10), and hypothalamus (N = 10). Full tissue-specific results are available in **Supplementary Table 2**. Major perturbagen were tyrosine kinase inhibitors including sorafenib, dasatinib, brivanib, and the novel therapeutic agent ursolic acid with multiple molecular targets. Drugs with gene targets that were found in the significant DEGs in our analysis in the AddNneuroMed cohort were utilized to create a drug-gene-network analysis (**Figure 5).**

### Predictors of Cognitive Decline: Modeling of MMSE Slope

Each of the three cognitive decline prediction models achieved significant accuracies per 5-fold cross-validation within the AddNeuroMed cohort (**Figure 3**). The clinical model achieved a Pearson’s *r* = 0.307 (95% CI [0.272–0.342], *z*-score = 7.46, *p*-value = 8.63×10 ¹) when predicting cognitive change over two years. Similar performance was found for the the omics model, reaching a Pearson’s correlation of 0.317 (95% CI [0.224–0.410], *z*-score = 7.73, *p*-value = 1.12×10 ¹). Integrating clinical and omics features into a single model (i.e., “unified model”) yielded the highest predictive accuracy, with a Pearson’s *r* = 0.393 (95% CI [0.311– 0.474], *z*-score = 9.75, *p*-value = 1.77×10 ²²).

**Figure 3.**
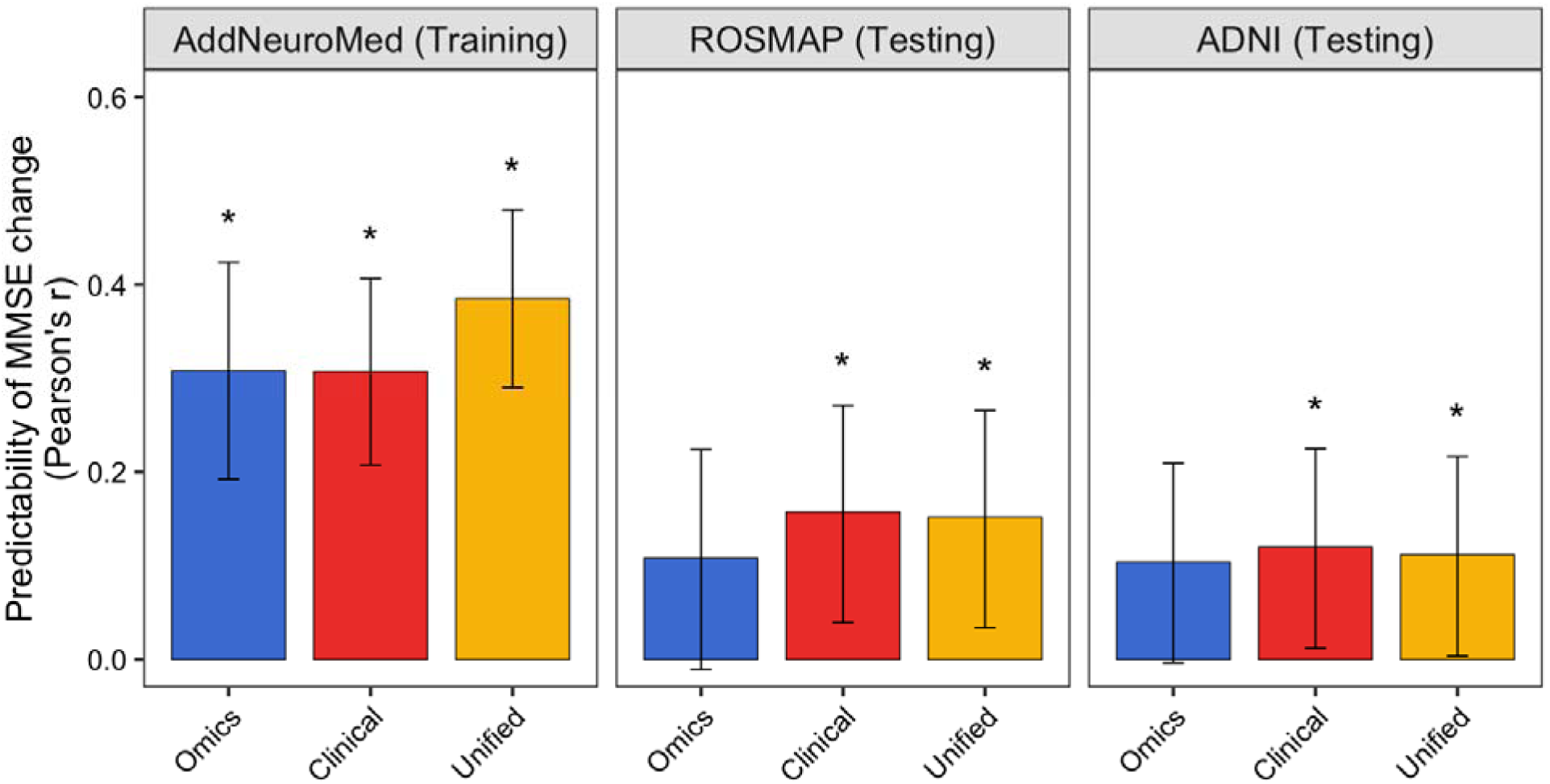
Accuracy of LASSO regression models tasked with predicting person-specific rate of MMSE change over a two-year assessment period.The panel specific to “AddNeuroMed” cohort shows the prediction accuracy of LASSO regression models from a single 5-fold cross-validation analysis. The 95% confidence interval is represented by the error bars. Asterisks (*) denote that prediction accuracy was statistically significant (*i.e*., *p* < 0.05). ROSMAP and ADNI cohorts served as independent testing samples of the LASSO regression models, optimized on data from the AddNeuroMed cohort.

We applied each prediction model to the ROSMAP cohort and found that the clinical and unified models achieved significant prediction performance indicating successful generalization (**Figure 3**). However, the accuracy of these models was comparatively modest relative to their performance when optimized in the AddNeuroMed cohort. Furthermore, the clinical model showed slightly better performance compared to the unified model when deployed on ROSMAP (clinical model: Pearson’s *r* = 0.157, 95% CI [0.039–0.271], *p*-value = 0.009; unified model: Pearson’s *r* = 0.152, 95% CI [0.034–0.266], *p*-value = 0.012), and the omics model did not generalize (Pearson’s *r* = 0.110, 95% CI [−0.009–0.225], *p*-value = 0.071). We again found that the clinical and unified models achieved statistically significant accuracy in the ADNI cohort (**Figure 3**) underscoring their external generalizability, albeit with modest overall accuracy (clinical model: Pearson’s *r* = 0.120, 95% CI [0.012–0.225], *p*-value = 0.029; unified model: Pearson’s *r* = 0.112, 95% CI [0.004–0.217], *p*-value = 0.042). The omics model was not statistically significant in the ADNI cohort (Pearson’s *r* = 0.104, 95% CI [−0.004–0.209], *p*-value = 0.059).

**Figure 4.**
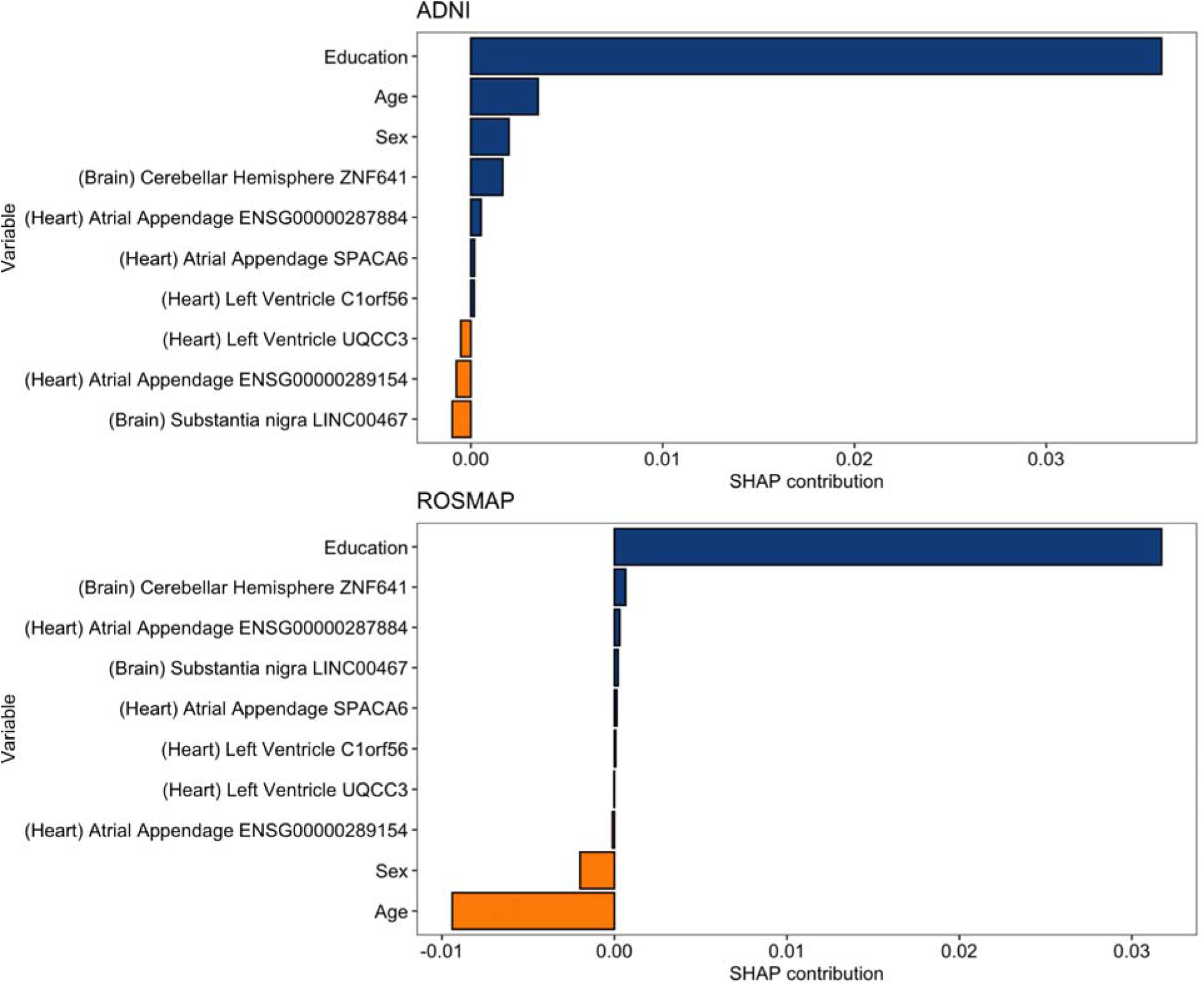
The predictive contributions of the 10 most informative features, according to the explainability SHAP algorithm, from the unified model that integrated brain-regional- and heart-imputed transcripts, age, sex, and years of education to predict person-specific decline in MMSE. Prediction contributions were computed based on the average contribution data from the participant in the ADNI and ROSMAP cohorts. Both cohorts were completely independent from the data collected from the AddNeuroMed used for model training.

### Model Explainability (SHAP Values)

SHAP explainability analysis quantified the mean contribution of each feature to prediction.The clinical models performed best in ROSMAP (*r* = 0.157) and ADNI (*r* = 0.120) in comparison to the omics and clinical model (ROSMAP *r* = 0.152; ADNI *r* = 0.112) and the omics-only model (ROSMAP *r* = 0.110; ADNI *r* = 0.103). Transcripts from the cerebellum, caudate and hypothalamus dominated the unified and omics-only models. The top contributors (designated via mean SHAP magnitude) in each cohort and model are summarized in Table 3 and Table 4.

**Figure 5.**
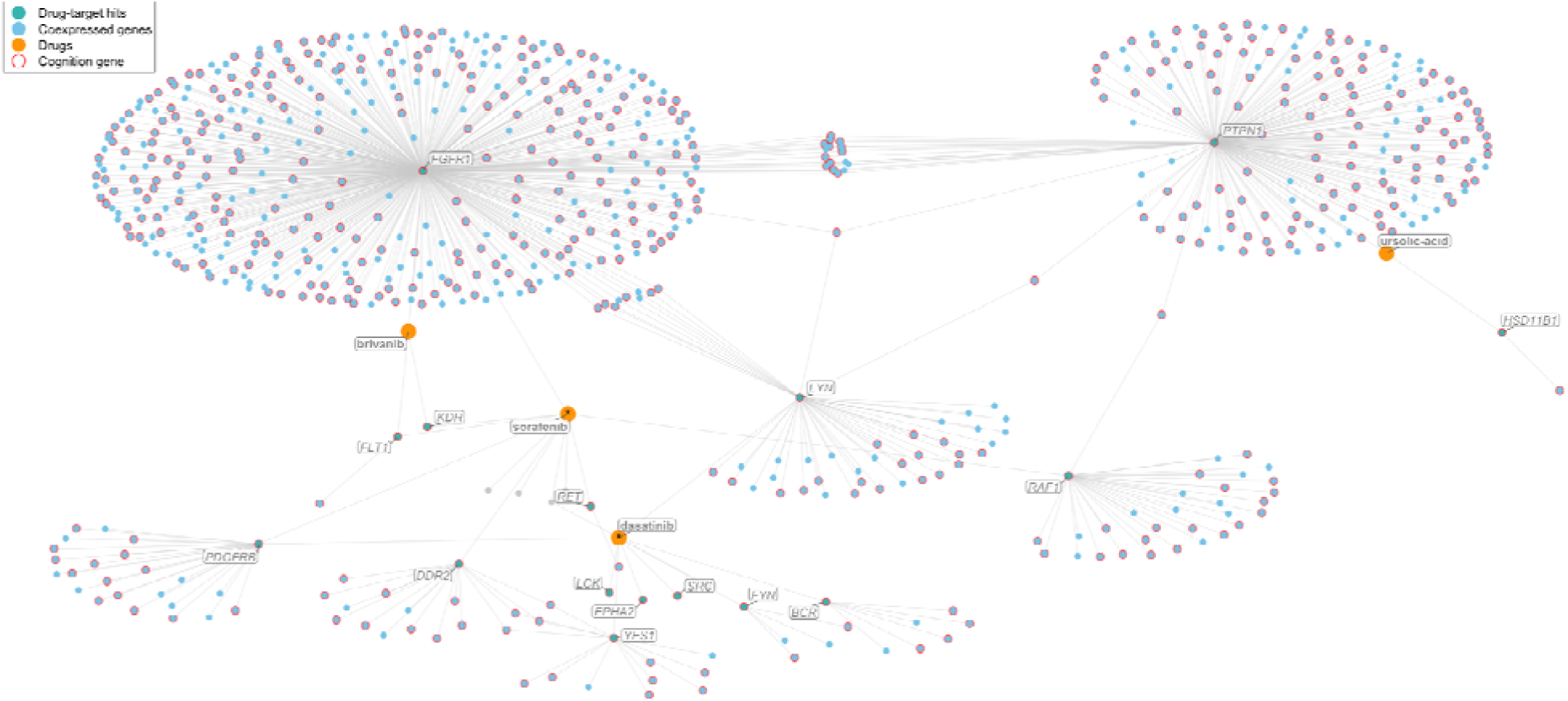
Drug–gene interaction network with significant pharmaceutical agents active on DEGs and imputed DEGs associated with cognitive changes in AddNeuroMed. Nodes represent drugs (orange), cognition-associated genes (red outline), and co-expressed genes (blue), with edges denoting either known drug–target relationships (drug–gene edges) or strong gene–gene co-expression (Pearson r ≥ 0.90). Cognition-associated genes were defined as those significantly associated with MMSE slope (FDR < 0.05) from linear models adjusting for age, sex, and education. Drug–target relationships were obtained from CMap compound annotations, and enrichment of drug–gene overlap was assessed using a hypergeometric framework, with significant compounds (FDR < 0.05) retained for visualization. Co-expression edges capture high-confidence transcriptional coupling between cognition-associated genes and the broader transcriptome. Asterisks indicate drugs predicted to be blood–brain barrier permeable. Network layout was generated using a force-directed algorithm (Fruchterman–Reingold), with hub-like structures reflecting drugs or genes with extensive connectivity. This visualization highlights candidate compounds whose known targets intersect cognition-related transcriptional signatures and their downstream co-expression neighborhoods.

**Table 3.**
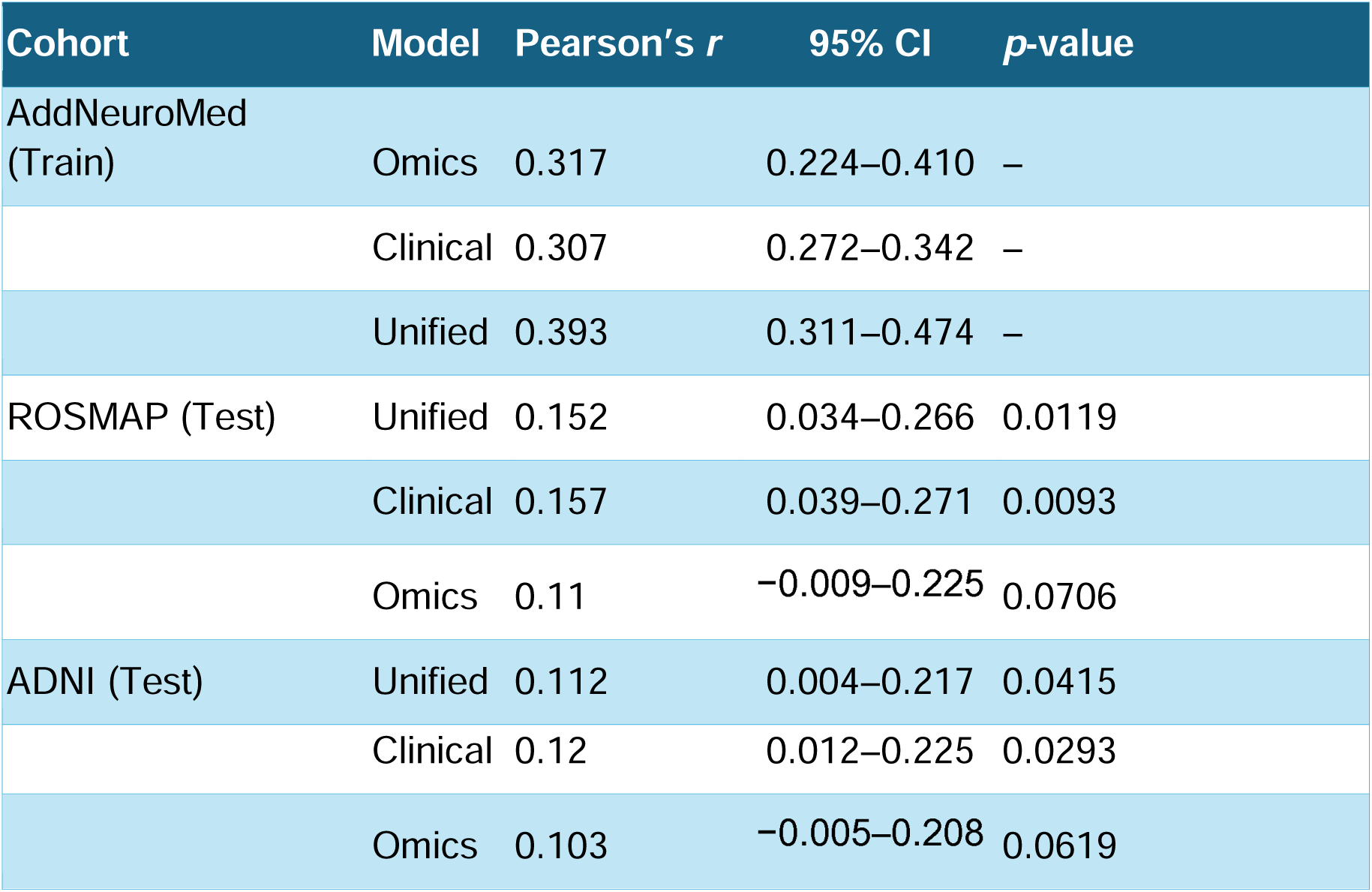
A performance summary for the three cognitive decline prediction models, evaluated through cross-validation (AddNeuroMed) and two independent test sets (ROSMAP and ADNI).

| Cohort | Model | Pearson's *r* | 95% CI | *p*-value |
| --- | --- | --- | --- | --- |
| AddNeuroMed (Train) | Omics | 0.317 | 0.224–0.410 | – |
|  | Clinical | 0.307 | 0.272–0.342 | – |
|  | Unified | 0.393 | 0.311–0.474 | – |
| ROSMAP (Test) | Unified | 0.152 | 0.034–0.266 | 0.0119 |
|  | Clinical | 0.157 | 0.039–0.271 | 0.0093 |
|  | Omics | 0.11 | –0.009–0.225 | 0.0706 |
| ADNI (Test) | Unified | 0.112 | 0.004–0.217 | 0.0415 |
|  | Clinical | 0.12 | 0.012–0.225 | 0.0293 |
|  | Omics | 0.103 | –0.005–0.208 | 0.0619 |

**Table 4.** Shapley analysis reveals the most influential features contributing to predictability of cognitive decline within the two test cohorts (ADNI and ROSMAP).

| Cohort | Model | Top Features (mean SHAP, sign) |
| --- | --- | --- |
| ADNI | Omics | (Brain) Cerebellar Hemisphere C3orf18 (+0.29); (Brain) Caudate IGFBP1 (−0.27); (Brain) Cerebellar Hemisphere ENSG00000274560 (+0.16); (Heart) LV SOD3 (+0.14); (Heart) LV FAM181B (−0.13) |
| ADNI | Unified | (Brain) Caudate IGFBP1 (−0.29); (Brain) Cerebellar Hemisphere C3orf18 (+0.23); (Heart) LV FAM181B (−0.15); (Brain) Cerebellar Hemisphere ENSG00000274560 (+0.13); (Heart) LV SOD3 (+0.11) |
| ADNI | Clinical | Education (+0.017); Sex (+0.016); Age (+0.012) |
| ROSMAP | Omics | (Brain) Cerebellar Hemisphere ENSG00000274560 (−0.35); (Brain) Cerebellar Hemisphere FAHD2B (−0.30); (Brain) Cerebellar Hemisphere SMIM10L2B (+0.30); (Brain) Cerebellar Hemisphere JAKMIP2 (+0.26); (Heart) Atrial Appendage RPS3AP26 (−0.26) |
| ROSMAP | Unified | (Brain) Cerebellar Hemisphere C3orf18 (+0.34); (Brain) Hypothalamus ERGIC1 (+0.32); (Brain) Cerebellar Hemisphere TRERF1 (−0.21); (Heart) Atrial Appendage ENSG00000289154 (−0.17); (Brain) Cerebellar Hemisphere SMIM10L2B (−0.15) |
| ROSMAP | Clinical | Age (−0.021); Education (+0.017); Sex (−0.010) |

## DISCUSSION

In this study, we applied the two imputation models *BrainGENIE* and *HeartGENIE* to predict gene expression across ten brain regions and two cardiac tissues, and utilized the predicted expression profiles and demographic and clinical characteristics to model longitudinal cognitive decline. Prediction of cognitive prmance can be useful for early identification of individuals at risk for rapid decline, patient prognosis, informing clinical trials, and planning of patient treatment (Petersen et al., 2018; Insel et al., 2020). Moreover, understanding the best biological predictors of cognitive decline, including specific transcripts, can provide mechanistic insights into the drivers of decline alongside actionable targets to interdict disease.

### Regional Associations

Associations between imputed brain- and cardiac-transcripts and MMSE 2-year slope were observed, with the greatest number of statistically significant transcripts (after Bonferroni correction) found in cerebellar hemisphere (225) and the two cardiac tissues—left ventricle (149 transcripts) and atrial appendage (113 transcripts). These results underscore the systemic cardiovascular changes associated with AD as well as cerebellar changes. We did not include participants’ MMSE scores at baseline in these downstream prediction models determined at their initial testing visit, as scores from the initial MMSE visit are factored in the calculation of slope of MMSE change (thus, conditioning on baseline MMSE would artificially enhance model performance

The correlation between cardiac transcripts and cognitive decline can be explained via 1) potential causal changes in cardiac function leading to impact on neural tissue and thus cognition, or 2) systemic vascular changes that are correlated between cardiac and brain tissue and cognitive decline. Multiple epidemiological studies have correlated cardiovascular diseases such as hypertension, atrial fibrillation, and heart failure with AD and cognitive decline, h**ig**hlighting cardiovascular dysfunction directly leading to cognitive changes in the case of stroke and hypoperfusion, and correlated changes such as coronary artery disease (Testai et al., 2024; Zhao et al., 2023; Iadecola et al., 2013; Gottesman et al., 2017). Vascular dysfunction can exacerbate amyloid and tau pathology, impair cerebral perfusion, and increase neuroinflammation. Several mechanisms have been proposed to explain this association. Chronic hypertension and small vessel disease can compromise the blood–brain barrier and cerebral autoregulation, increasing susceptibility to amyloid deposition and white matter injury (Iturria-Medina et al., 2016). Cardiac dysfunction reduces cerebral perfusion, producing chronic hypoxia that accelerates tau pathology and neuronal loss (de la Torre, 2012). Systemic inflammation exacerbated via cardiovascular disease may also propagate neuroinflammatory cascades implicated in AD pathogenesis (Venkat et al., 2015). Together, these pathways highlight multiple routes through which molecular changes in heart tissue can drive or exacerbate neurodegeneration. The identification of cardiac transcripts associated with MMSE decline raises the possibility that transcriptional changes in peripheral organs may act as systemic correlated indicators or potential contributors to neurodegenerative processes.

The cerebellar hemisphere had the greatest number of significantly associated transcripts. Involved in motor coordination, the cerebellum has been implicated in cognitive and affective processes, and its changes in AD have been highlighted in neuroimaging and postmortem studies. Imaging studies have shown cerebellar atrophy in AD that correlates with memory impairment and disease severity (Thomann et al., 2008; Guo et al., 2016). Moreover, disrupted cerebello-cortical circuits have been shown to be correlated to cognitive symptoms (Jacobs et al., 2018). Postmortem transcriptomic studies have shown dysregulation in synaptic and immune pathways in the cerebellum (Miller et al., 2013; Wan et al., 2020). The findings presented here offer further support for cerebellar changes associated with AD progression.

### Pathway Enrichment Across Tissues

Regional and pathway-based patterns emerged in the root pathway analysis of the transcripts associated with cognitive decline. Widespread downregulation in the heart tissues (right atrial appendage and anterior left ventricle) and substantia nigra, and widespread upregulation of pathways in the cerebellar hemisphere were notable regional findings. Postmortem studies of cognitive decline have validated changes in substantia nigra and the cerebellar hemisphere tissues (Ferrari et al., 2024) (Gaunt et al., 2023). Moreover, gene expression changes in the atrial appendage have been associated with amyloidopathy (Hohman et al., 2017).

Downregulated pathways across tissues included metabolism, protein metabolism, cellular response to stimuli and disease andupregulated pathways across tissues included signal transduction, immune, chromatin organization and disease. Postmortem studies of Alzheimer’s disease have replicated widespread regional downregulation in metabolism, metabolism of proteins, cellular response to stimuli and disease as well as upregulation in signal transduction and chromatin organization (Hou et al., 2025).

Metabolic changes in the setting of dementia can be categorized into dysregulation in glucose and mitochondrial metabolism (Bano et al., 2023), oxidative stress and calcium homeostasis (Butterfield & Halliwell et al., 2019), and lipid (yin et al., 2023) and protein metabolism (Soto et al., 2018). Whether these changes are upstream or downstream of tissue atrophy and damage is a matter of extensive debate. Proteinopathy has been investigated as one of the leading associated and potentially causative dysregulations in various types of dementia, with amyloid, TDP43, tauopathy, alpha synuclein axis of proteinopathies involved in Alzheimer’s DIsease, Parksnson’s DIsease, frontotemporal dementia (Soto et al.,2018)

Given our current approach, access to longitudinal blood data would allow us to better understand the corresponding molecular changes in different stages of pathological stages as they evolve, and potentially increase our nuance of cause and effect in dementia biology.

### Drug Perturbation Signatures (CMap)

The current state of drug repurposing and discovery in dementia is focused on addressing the posited root causes such as amyloidopathy in AD. Nevertheless, most recent agents tackling amyloidopathy via antibody mediated clearance have failed to demonstrate clinically significant disease course modification (van Dyck et al., 2023; Sims et al., 2023). Examples of agents currently under investigation include NAD+ restoring compounds with promise in animal (Xiong et al., 2024), anti-sense RNA agents are under investigation for tauopathy (Mummary et al., 2023), and metformin as a metabolic regulator able to change AD pathological development (Luchsinger et al., 2025). Given the current landscape, there are major opportunities to expand the potential space of compounds and mechanisms that could be investigated and studies such as this extend the currently discussed putative agents.

The drug repurposing analysis found molecules that show positive and negative correlations with cognitive decline signature in different tissues. The findings highlight candidate compounds for further investigation. Our drug-genenetwork analysis identified several perturbagens having known pharmacological interactions with cognition-related transcripts found in this study including:sorafenib, dasatinib, brivanib and ursolic acid. Sorafenib, dasatinib, brivanib are kinase inhibitors acting on tyrosine kinases with downstream inhibition of MAPK/RAF signaling and are currently used for various cancers(U.S. Food and Drug Administration, 2020, 2024; Jones et al., 2019). Urosilic acid is a novel multi-target drug with activity in the NF-κB, PI3K/Akt, MAPK, and STAT3 pathways,decreasing inflammation and increasing survival signaling (Khwaza et al., 2024).

Inflammatory and pro-survival pathways have been implicated in dementia and suggested as potential targetable pathways. Ahmed et al., 2020 discuss the opportunity for MAPK targeting Alzheimer’s disease therapeutic development. STAT3 signaling is linked to neuroinflammation in AD pathology. Wen et al., 2024 have pointed to STAT3 signaling as a potential targetable pathway in AD. PI3K/AKT signaling is involved in cellular survival, metabolic regulation, and synaptic plasticity and has been implicated in AD pathological development (Li et al., 2024). Pan et al., 2024 and Long et al., 2021 have posited this pathway as a potential therapeutic target for AD as well.

### Predictive Modeling and Explainability

Modelling showed a modest increase in cognitive decline prediction performance with the addition of imputed transcriptomic data to clinical and demographic data. The unified models, integrating age, sex, education, and imputed expression explained an additional 2.5% of variance in pace of cognitive decline, outperforming the other models in cross-validation (AddNeuroMed). The unified model also outperformed the demographic characteristics-models in the external cohorts (ROSMAP and ADNI), indicating generalizability of these findings. Explainability analyses implicated demographic characteristics as the main drivers of performance in the unified model. Yet, cerebellar and cardiac imputed transcripts provided predictive performance beyond known predictors of cognitive decline, and ranked among the top predictors in the unified model as demonstrated in Table 4. These findings indicate the potential utility of brain- and heart-predicted transcripts identifying novel molecular signatures associated with cognitive decline.uture studies including larger sample sizes and more consistent site and data-collection methodologies may improve prediction of cognitive decline with these promising new markers.

### Limitations and Future Directions

This study has multiple limitations. First, our transcriptome-imputation models recover brain- and heart-specific gene-expression signatures, yet they are intended as a supplement to, but not substitute for, direct tissue measurements.

Second, our study focused on the slope of MMSE change within a two-year follow-up window. Given the typical annual rate of change in memory in dementia patients,^8^ a two-year window affords sufficient time for memory changes to accumulate and become clinically interpretable. It also provides better sensitivity to detect real memory changes, avoiding the error introduced by “practice effects” from repeat assessments. A longer follow-up window may help to reveal novel signatures in brain and heart gene-expression that bear on cognitive performance, yet the limited sample size available beyond the two-year follow-up window would have left our study underpowered to detect robust associations.

Third, blood RNAs were not serially sampled from the dementia cohorts included in this study, thus we cannot distinguish correlative from causally linked drivers of cognitive decline. Obtaining longitudinal blood RNAs in future patient cohorts may help reveal brain- and cardiac-specific gene-expression trajectories contributing to rapid and slow cognitive decline among individuals with dementia.

Lastly, cognitive decline was better predicted in the cohort used for model optimization (AddNeuroMed) compared to the cohorts in which the models were externally tested. Statistically significant external generalizability is critical for demonstrating feasibility and viability, which was achieved in this study. Furthermore, the models were able to capture reproducible signals anchored to cognitive decline. However, their decline in accuracy and overall modest performance should be interpreted with caution. Several factors may explain this attenuation of model performance. The external cohorts (ROSMAP and ADNI) used for evaluating model generalizability differ from our training set (AddNeuroMed) on key technical and clinical characteristics, including the profiling methodologies for measuring blood RNAs and the composition of normal aging versus dementia cases. Accordingly, our models provide a conservative estimate of the *lower bound* of model accuracy, which may be improved upon with better matching of cohorts and further control against overfitting.

## SUMMARY

Our findings offer novel insight into transcripts correlated with cognitive decline in different tissues. Notably, cardiac tissue changes showcase changes in peripheral biology that are correlated with cognitive decline and warrant further study into potential shared biology. Our models have indicated the utility of added transcriptomic data in predicting cognitive decline alongside clinical and demographic characteristics. Our drug correlation analysis further suggests positive and negative correlations that could set the ground for further drug repurposing studies. This study offers a novel application of blood-based imputation models for the discovery of transcriptomic changes associated with cognitive decline.

## DATA AVAILABILITY

We obtained access to controlled data for donors in ROSMAP and AddNeuroMed upon approval *via* Synapse (ROSMAP: https://www.synapse.org/Synapse:syn22024496; AddNeuroMed: https://www.synapse.org/Synapse:syn22252881) and from the ADNI website at https://ida.loni.usc.edu/. Individual-level data are not available to be shared.

## CODE AVAILABILITY

All analyses and data visualization were performed using custom *R* scripts. The scripts can be provided upon written request.

## FUNDING ACKNOWLEDGEMENTS

J.L.H. is supported by grants from the U.S. National Institutes of Health (R21MH126494, R01NS128535), the Central New York Community Fund, and the NARSAD: The Brain & Behavior Research Foundation (2020 Young Investigator Award). S.J.G. is supported by grants from the U.S. National Institutes of Health (R01AG064955 and R21MH126494), the U.S. National Science Foundation, the Sidney R. Baer, Jr. Foundation, and NARSAD: The Brain & Behavior Research Foundation. S.V.F. is supported by the European Union’s Horizon 2020 research and innovation programme under grant agreement 965381; NIH/NIMH grants U01AR076092, R01MH116037, 1R01NS128535, R01MH131685, 1R01MH130899, U01MH135970, the Upstate Foundation, Corium Pharmaceuticals, Tris Pharmaceuticals, and Supernus Pharmaceutical Company. A.J.G. is supported by a grant from the U.S. National Institutes of Health (K08HL168205). N.R.T. is supported by a grant from the U.S. National Institutes of Health (R01HL170051).

